# Trends in Prescriptions of Lithium and Other Medications for Patients with Bipolar Disorder in Office-based Practices in the United States: 1996-2015

**DOI:** 10.1101/2020.01.18.20018028

**Authors:** Yian Lin, Ramin Mojtabai, Fernando S Goes, Peter P Zandi

## Abstract

**Background:** Studies have shown that rates of lithium use for bipolar disorder (BPD) in the United States declined through the 1990s as other mood stabilizing anticonvulsants and second-generation antipsychotics (SGAs) became more popular. We examined recent prescribing trends of medications for BPD over the past two decades.

**Methods:** Twenty years of data (1996-2015) from the National Ambulatory Medical Care Survey (NAMCS) were used. Weighted percentages of prescriptions of lithium, anticonvulsants, SGAs and antidepressants were calculated over two-year intervals. Logistic regression was used to examine factors related to polytherapy.

**Results:** Prescriptions of lithium declined from 38.1% (95%CI: 29.8% - 46.3%) in 1996-97 to 14.3% (95%CI: 10.6% - 18.1%) in 2006-07 and has remained stable since. During this time, prescriptions of SGAs more than doubled. SGAs and/or anticonvulsants were prescribed in 78.6% (95%CI: 73.0% - 84.2%) of BPD visits in 2014-2015. Polytherapy increased by approximately 4% every two years and in 2014-15 occurred in over 35% of BPD visits. Antidepressants were prescribed in 40-50% of BPD visits, but their prescriptions without other mood stabilizers decreased from 18.2% (95%CI: 11.7% - 24.8%) in 1998-99 to 5.8% (95%CI: 3.0% - 8.6%) in 2014-15.

**Limitations:** The sample had limited power to study the effect of individual medications or the potential for differing effects in certain subgroups of patients.

**Conclusions:** This study further documents the declining prescriptions of lithium for BPD, and corresponding increase in prescriptions of anticonvulsants and SGAs, despite the fact that lithium is typically recommended as a first line therapy for BPD.

## Introduction

Bipolar disorder (BPD) is a severe and often disabling psychiatric disorder that affects approximately 1-4.4% of the world’s population (Merikangas et al., 2007; Merikangas et al., 2011). BPD is classically described as having a course of illness that alternates between extreme mood states of mania and depression and is characterized by episodic relapses along with longstanding disability (Nierenberg et al., 2013). BPD is a major cause of hospitalizations and health care expenditures, and is also associated with an increased risk of suicide (Chesney, Goodwin, & Fazel, 2014). Although a number of medications have been approved for treatment of acute episodes or for maintenance treatment of BPD, lithium, the first medication shown to be a mood stabilizer in 1949 (Cade, 1949), remains a first line therapy for BPD (Severus, Schaaff, & Moller, 2012). However, it is acknowledged that the efficacy of lithium varies widely, with about 30% of treated patients showing only partial response and another 25% having no response (Hou et al., 2016). In addition, lithium use is associated with certain serious side effects (McKnight et al., 2012), requiring careful clinical management due to its narrow therapeutic index (Okusa & Crystal, 1994).

This has led to the increasing use of other medications that also have demonstrated efficacy in BPD, including certain anticonvulsants, second-generation antipsychotics (SGAs), and antidepressants (Geddes, Calabrese, & Goodwin, 2009; M. J. Gitlin, 2018; Lindstrom, Lindstrom, Nilsson, & Hoistad, 2017; Weisler, Cutler, Ballenger, Post, & Ketter, 2006). However, anticonvulsants and antipsychotics may be less effective than lithium in preventing the recurrence of mood episodes or in reducing the overall risk of suicide (Caley, Perriello, & Golden, 2018; Kessing, Sondergard, Kvist, & Andersen, 2005). In addition, the appropriate clinical use of antidepressants in BPD remains uncertain (M. J. Gitlin, 2018; Sachs et al., 2007).

Nevertheless, there has been an increase in polytherapy with combinations of these medications. There is some evidence that certain combinations of medications may be superior to monotherapy for treating manic episodes, but continued polytherapy after the manic phases resolve remains controversial (Geoffroy, Etain, Henry, & Bellivier, 2012).

The trends in the use of various medications for treatment of BPD have been documented by several studies in European countries (Bramness, Weitoft, & Hallas, 2009; Castells et al., 2006; Hayes et al., 2011; Wilting, Souverein, Nolen, Egberts, & Heerdink, 2008). In the US, trends in prescriptions of lithium and other medications for treatment of BPD have been examined through the 1990s and early 2000s using data from the National Ambulatory Medical Care Survey (NAMCS) and Kaiser Permanente, Northern California health system (Blanco, Laje, Olfson, Marcus, & Pincus, 2002; Hunkeler et al., 2005; Moreno et al., 2007). However, there is little national level data on trends over the past 20 years. Data from a single tertiary care clinic suggested a decrease in lithium usage (Hooshmand et al., 2014); however, it is unclear whether this trend is generalizable to other settings.

In this study, we examined trends in prescriptions of lithium and other medications for BPD in a representative sample across the US over a period of 20 years. We used data from NAMCS, a well-characterized national survey of US office-based practices of physicians from different specialties, conducted annually across a broad range of office settings. Specifically, we examined trends in prescriptions of lithium, mood-stabilizing anticonvulsants, SGAs, and antidepressants in visits with a diagnosis of BPD. The results of our study provide useful data to characterize the changing patterns in the management of BPD in the US over the past two decades.

## Methods

### Study Data

For the current analysis, we used 20 years of data from the NAMCS between 1996 to 2015 to summarize and analyze recent trends in prescriptions of lithium and other medications for BPD. The NAMCS has been conducted annually since 1989 utilizing a multistage probability design in which a sample of patient visits are randomly selected with a pre-specified probability from within certain physician practices, which in turn are randomly selected with a pre-specified probability from within primary sampling units (PSUs) across the country (CDC, 2015a). Data are obtained on patient, visit, and physician practice characteristics (CDC, 2017). The current analysis was considered exempt from human subject research, because the data were publicly available and de-identified.

We identified all visits in which the patient had a diagnosis of BPD as indicated by a primary, secondary, or tertiary diagnosis with one of the following ICD-9 codes: 296.00-296.06, 296.10-296.16, 296.40-296.46, 296.50-296.56, 296.60-296.66, 296.7, 296.80-296.81, and 296.89. While the number of diagnoses that could be recorded per visit increased over the years, we extracted the first three diagnoses for each encounter to be consistent across all years.

Medications prescribed at each visit were coded using the National Drug Code Directory (NDCD) prior to 2006 and the Multum drug dictionary starting in 2006 (CDC, 2015b). While the number of medications that could be recorded per visit in NAMCS has increased over the years, we extracted the first six medications for each encounter to be consistent across all years.

We categorized medications prescribed for BPD based on their NDCD or Multum codes into the following classes: lithium, anticonvulsants used as mood stabilizers (valproic acid [divalproex sodium], carbamazepine, lamotrigine, and oxcarbazepine), second-generation antipsychotics (SGAs) (aripiprazole, asenapine maleate, clozapine, iloperidone, lurasidone, olanzapine, olanzapine/fluoxetine, paliperidone, quetiapine, risperidone, and ziprasidone), and antidepressants (clomipramine, amoxapine, nortriptyline, citalopram hydrobromide, duloxetine, trazodone HCl, amitriptyline, venlafaxine HCl, selegiline, perphenazine/amitriptyline, fluvoxamine maleate, escitalopram oxalate, chlordiazepoxide/amitriptyline, maprotiline, isocarboxazid, phenelzine sulfate, nefazodone HCl, desipramine HCl, tranylcypromine sulfate, paroxetine HCl, and paroxetine mesylate). We considered patients as receiving “no medication treatment” for BPD at a visit if they were not prescribed any of the above medications at that visit.

Patient characteristics included sex, race (white, black, others), and age (adults, children); patients under the age of 18 were considered children. The insurance type used to cover visits was classified as either private insurance, Medicaid/Children’s Health Insurance Program (CHIP), Medicare, self-pay, or others. The visit type was defined as either “psychiatry” if the visit was to a psychiatrist, or “others” for all other office-based physicians.

### Data Analysis

Visits in which lithium was prescribed were compared to those in which other or no mood stabilizer medications were prescribed with regard to patient and visit characteristics using contingency tables and chi-square tests. Trends over time in the use of the different medication classes for BPD were described as weighted percentages calculated from the number of visits in which a particular category of medication was prescribed out of the total number of BPD visits in a given year. We further examined trends in prescriptions of the different mood stabilizer classes within important patient sub-groups, including by BPD subtypes and age, as wells as trends in prescriptions of the top four most prescribed anticonvulsants and SGAs. Next, we used logistic regression to examine the association between patient and visit characteristics and polytherapy of mood stabilizer medications (i.e., lithium, anticonvulsants and/or SGAs). In the logistic regression analyses, we included only BPD visits in which medications from any of the mood stabilizer classes were prescribed. All analyses were adjusted for visit weights, clustering, and stratification of data using design elements provided by the National Center for Health Statistics that allow generalization of the NAMCS data to the US population. In order to obtain more stable estimates of the prescription rates, we combined visit data over two-year intervals. All analyses were conducted in RStudio (Version 1.1.447).

## Results

### Sample Characteristics

Between 1996 and 2015, the NAMCS included data on 645,784 visits to office-based physicians; of these, 5,400 (0.6%) visits involved patients who had a BPD diagnosis, including approximately 47% with bipolar I disorder, 13% with bipolar II disorder, and 40% with bipolar disorder not otherwise specified. **Table 1** summarizes the characteristics of these BPD visits and compares them by whether lithium, other mood stabilizers (anticonvulsants and/or SGAs), or no mood stabilizers were prescribed. The majority of visits involved white adults, and just over 60% were with females. Over three-quarters of the visits were with psychiatrists, and over 40% were covered by private insurance, while another 35% were covered by Medicare and/or Medicaid. Lithium was prescribed in approximately 17% of BPD visits, other mood stabilizers (anticonvulsants and/or SGAs) were prescribed in 53%, and no mood stabilizers were prescribed in 30%. There were significant differences in race, age, visit type, insurance type, and prescriptions of antidepressants across the 3 treatment groups (all p-values < 0.0001).

**Table 1.** Characteristics of visits with a BPD diagnosis by mood stabilizer treatment ^*i*^

|  | Bipolar<br>(n = 5,400) | Lithium<sup>ii</sup><br>(n = 925) | Other Mood<br>Stabilizers<sup>iii</sup><br>(n = 2,864) | No Mood<br>Stabilizers<br>(n = 1,611) | *p*-value<sup>iv</sup> |
| --- | --- | --- | --- | --- | --- |
| *Sex* |  |  |  |  | 0.1475 |
| Female | 61.6 | 58.8 | 62.0 | 62.5 |  |
| Male | 38.4 | 41.2 | 38.0 | 37.5 |  |
| *Race* |  |  |  |  | < 0.0001 |
| White | 88.1 | 91.5 | 87.1 | 88.4 |  |
| Black | 7.9 | 3.7 | 8.6 | 8.5 |  |
| Others | 4.0 | 4.8 | 4.2 | 3.1 |  |
| *Age* |  |  |  |  | < 0.0001 |
| Adults (≥ 18 years) | 91.6 | 94.4 | 89.2 | 94.1 | |
| Children | 8.4 | 5.6 | 10.8 | 5.9 |  |
| *Visit Type* |  |  |  |  | < 0.0001 |
| Psychiatry | 76.8 | 83.0 | 83.5 | 61.2 |  |
| Others | 23.2 | 17.0 | 16.5 | 38.8 |  |
| *Insurance Type* |  |  |  |  | < 0.0001 |
| Private Insurance | 43.9 | 43.9 | 45.2 | 41.7 |  |
| Medicaid,CHIP | 17.3 | 11.8 | 18.9 | 17.5 |  |
| Medicare | 17.6 | 19.3 | 15.8 | 19.8 |  |
| Self-pay | 13.6 | 16.6 | 12.9 | 13.2 |  |
| Others | 7.6 | 8.3 | 7.2 | 7.8 |  |
| *Antidepressants* |  |  |  |  | < 0.0001 |
| Yes | 46.9 | 42.3 | 53.7 | 37.4 |  |
| No | 53.1 | 57.7 | 46.3 | 62.6 |  |

<sup>i</sup> All numbers shown in the table are weighted percentages (see **Methods**).
<sup>ii</sup> Among visits being prescribe lithium, 444 were prescribed lithium monotherapy (as the only mood stabilizer), 135 were concomitantly prescribed anticonvulsants (but no SGAs), 238 SGAs (but no anticonvulsants), and 108 both.
<sup>iii</sup> Among visits being prescribed non-lithium mood stabilizers, 1,085 were prescribed anticonvulsants as the only mood stabilizers, and 1,036 SGAs. 743 were concomitantly prescribed anticonvulsants and SGAs.
<sup>iv</sup> P-values are from Chi-squared tests comparing percentages across the treatment groups (lithium; other mood stabilizers; and no mood stabilizers).

Prescriptions of any mood stabilizers were more likely to occur at psychiatry visits. Lithium prescriptions were more likely to involve white adult patients who were self-paying and not on Medicaid. Compared to lithium, prescriptions of other mood stabilizers were more likely among children and to be accompanied with prescriptions of antidepressants.

### Trends in Mood Stabilizer Classes

The percentage of BPD visits among all visits steadily increased over the study period from 0.3% (95% CI: 0.2% - 0.4%) in 1996-97 to 0.8% (95% CI: 0.6% - 1.0%) in 2014-15. Among the BPD visits, the percentage in which any mood stabilizer was prescribed remained stable at approximately 70%, until 2014-15, when there was an increase to 82.1% (95% CI: 77.1% - 87.1%). However, during this period there was a consistent decline in prescriptions of lithium (**Figure 1**). The percentage decreased from 38.1% (95% CI: 29.8% - 46.3%) in 1996-97 to 14.3% (95% CI: 10.6% - 18.1%) in 2006-07, and it remained steady at approximately 14% from 2008-09 on. During this period, there was a parallel increase in prescriptions of SGAs, which more than doubled from 19.2% (95% CI: 12.4% - 26.0%) in 1996-97 to 48.0% (95% CI: 42.2% - 53.9%) in 2008-09 and reached a peak of 52.2% (95% CI: 43.9% - 60.5%) in 2014-15. Prescriptions of anticonvulsants for BPD remained generally stable at around 40%, with a low of 31.0% (95% CI: 23.9% - 38.1%) in 1996-97 and a recent peak of 49.8% (95% CI: 43.2% - 56.4%) in 2014-15. Overall, by 2014-15, other mood stabilizers (anticonvulsants and/or SGAs) were prescribed in 78.6% (95% CI: 73.0% - 84.2%) of BPD visits.

**Figure 1.**
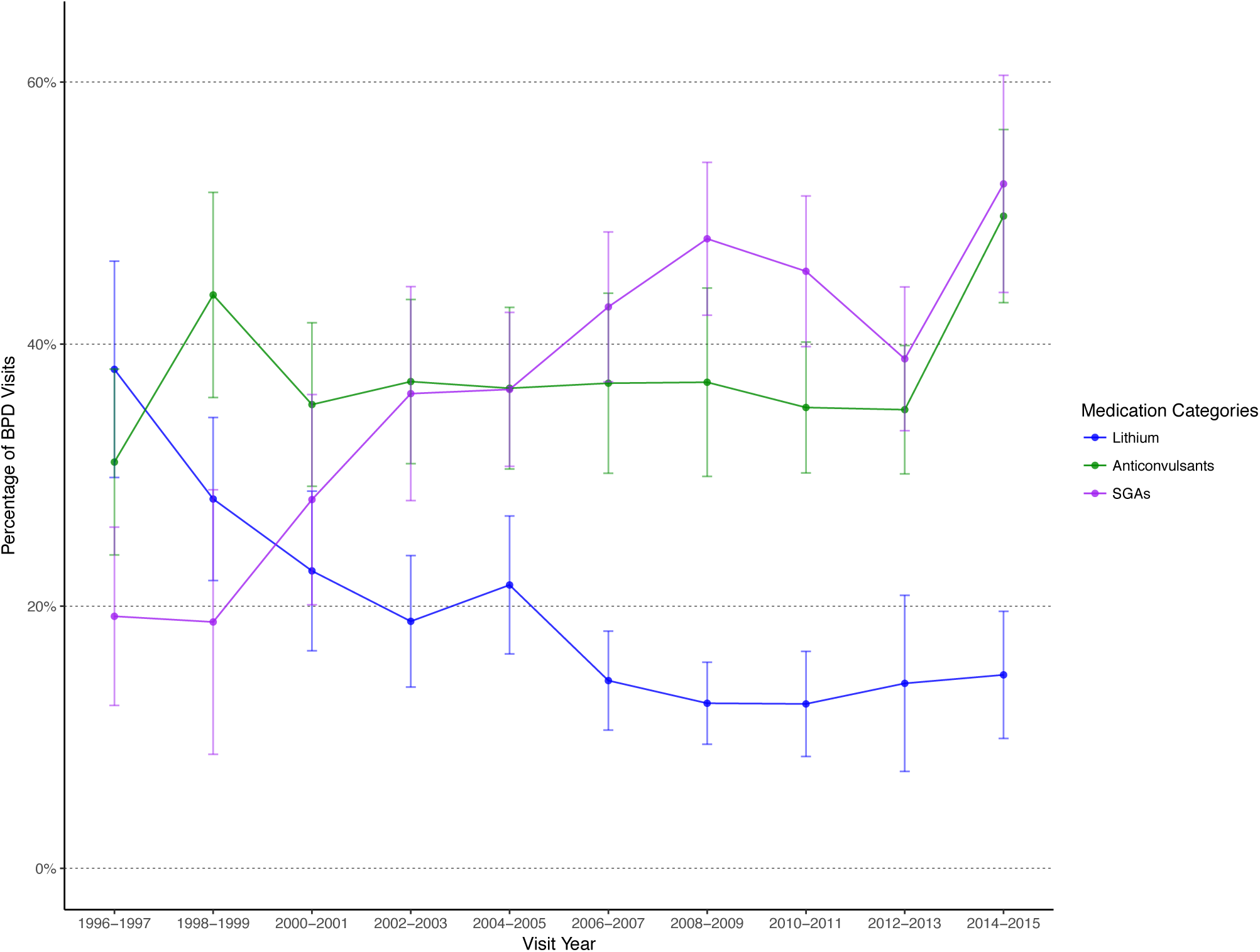
Trends between 1996 and 2015 in percentage of BPD visits for different categories of mood stabilizer medications for BPD ^*i*^. *i* Error bars show for each weighted estimate of the percentage of BPD visits the 95% confidence interval.

### Trends by Cohort Sub-Groups

Trends in prescriptions of different mood stabilizer classes were generally similar for patients with different BPD subtypes, except for a somewhat greater overall prescriptions of lithium for bipolar I disorder compared to other bipolar diagnoses (**Supplementary Figure 1**). By the end of 2014-15, 19.5% (95% CI: 11.0% - 27.9%) of visits for patients with bipolar I disorder were prescribed lithium compared to only 11.6% (95% CI: 7.0% - 16.2%) of visits for patients with other bipolar diagnoses.

Trends in prescriptions of the different classes of mood stabilizers were also generally similar when comparing adults with children (age < 18 years old), except for two noteworthy differences. First there was greater variability in estimates over time for children due to the smaller sample size, and second, there was a notable difference in SGAs prescriptions (**Supplementary Figure 2**). The decline in prescriptions of SGAs seen after 2008-09 occurred in both adults and children, but was more precipitous in children, where prescriptions of SGAs dropped from a high of 70.1% (95% CI: 55.0% - 86.7%) in 2008-09 to 43.2% (95% CI: 26.1% - 60.3%) in 2012-13. However, prescriptions of SGAs in children rebounded in 2014-15 to a high of 78.4% (95% CI: 50.2% - 100%), although given the wide confidence intervals, it is unclear if this reflects a genuine change or is due to unstable estimates from a small sample size.

### Trends in Specific Anticonvulsants and SGAs

Trends in prescriptions of the top four most prescribed anticonvulsants and SGAs between 1996-2015 are shown in **Figure 2**. Among anticonvulsants, there was a notable decline in prescriptions of valproic acid/divalproex sodium coupled with an increase in lamotrigine. The percentage of BPD visits with prescriptions of valproic acid/divalproex sodium peaked in 1998-99 at almost 35%, but then declined to around 14% in 2014-15. Conversely, as of 2006-07 lamotrigine became the most widely prescribed anticonvulsant and was prescribed in approximately 28% of BPD visits in 2014-15. Among SGAs, there was a steady increase in the percentage of BPD visits with prescriptions of quetiapine and aripiprazole since their introduction. Beginning in 2004-05, quetiapine became the most widely prescribed SGA, being prescribed in 22.0% (95% CI: 15.3% - 28.7%) of BPD visits in 2014-15. Since 2008-09, aripiprazole has been the second most widely prescribed SGA and was prescribed in 12.3% (95% CI: 7.4% - 17.2%) of BPD visits in 2014-15.

**Figure 2.**
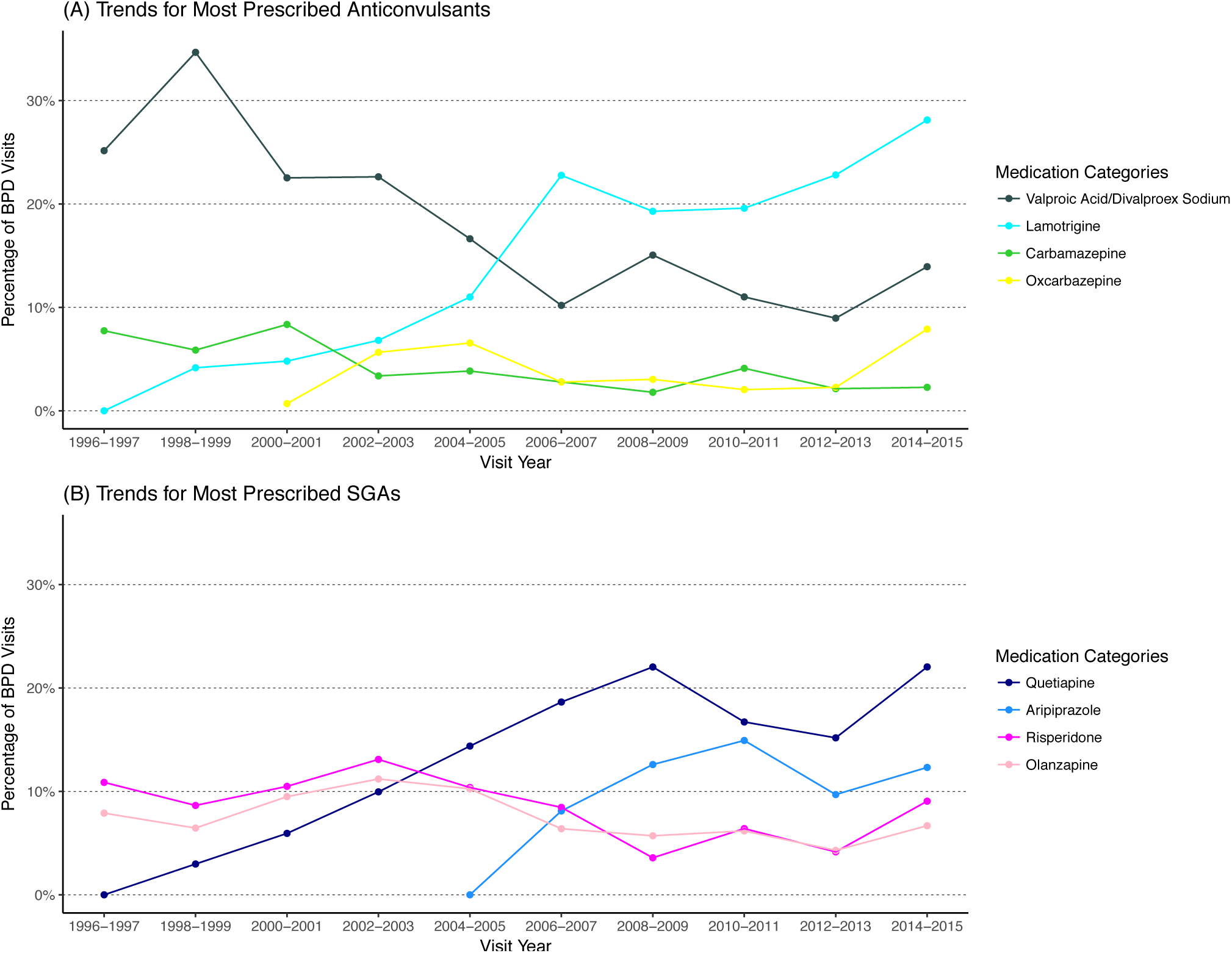
Percentage of BPD visits over time of specific anticonvulsants and SGAs ^*i, ii*^. *i* Error bars are not shown due to smaller sample sizes of the stratified results. *ii* Oxcarbazepine and aripiprazole entered the US market later than 1996, which explains why oxcarbazepine first appears in the NAMCS data in 2000-01, and aripiprazole in 2004-05.

### Trends in Polytherapy

The use of multiple medications to treat mood in BPD has steadily increased since 1996-97. In 1996-97, two or more mood stabilizers were prescribed in only 20.0% (95% CI: 13.1% - 26.8%) of BPD visits; however, by 2014-15 the percentage had almost doubled to 37.0% (95% CI: 27.8% - 46.1%) (**Figure 3**). Among BPD visits in which any mood stabilizer treatment was prescribed, the odds of polytherapy with mood stabilizers increased by 1.04 times (95% CI: 0.99 - 1.09) every two years, after controlling for sex, race, age, visit type, and insurance type. Of these other covariates, the provider type was significantly associated with increased polytherapy, the odds being 3.25 (95% CI: 2.21 - 4.79) times greater in psychiatric versus non-psychiatric visits.

**Figure 3.**
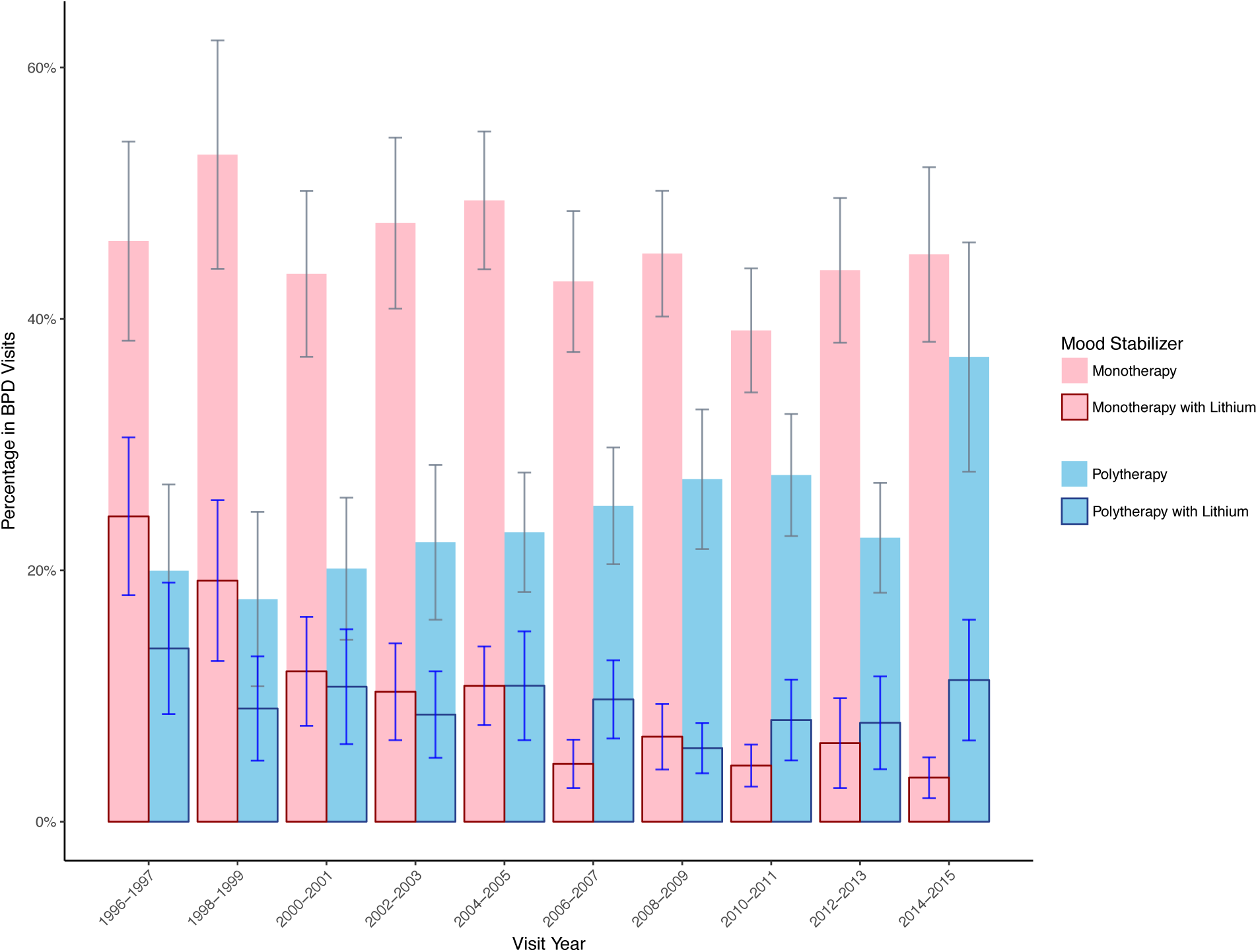
Trends in monotherapy vs. polytherapy of mood stabilizer medications for BPD^*i*^. *i* Error bars show for each weighted estimate of the percentage of BPD visits the 95% confidence interval.

Despite the general increase in polytherapy of mood stabilizers over the past two decades, prescriptions of lithium concomitantly with other mood stabilizers declined somewhat from a high of 13.8% (95% CI: 8.6% - 19.0%) in 1996-97 to 11.3% (95% CI: 6.5% - 16.1%) in 2014-15. Over this same time, prescriptions of lithium monotherapy decreased even more dramatically from 24.3% (95% CI: 18.0% - 30.6%) in 1996-97 to only 3.5% of BPD visits (95% CI: 1.9% - 5.1%) in 2014-15.

### Trends in Antidepressants

Antidepressants were consistently prescribed in approximately 40-50% of BPD visits (**Figure 4**). Moreover, the percentage of visits in which antidepressants were prescribed by themselves without any mood stabilizers decreased from 18.2% (95% CI: 11.7% - 24.8%) in 1998-99 to 5.8% (95% CI: 3.0% - 8.6%) in 2014-15. At the same time, the percentage of visits in which antidepressants were prescribed with mood stabilizer(s) steadily increased from 26.3% (95% CI: 20.2% - 32.3%) in 1996-97 to 45.8% (95% CI: 38.2% - 53.4%) in 2014-15. Prescriptions of antidepressants were generally lower in children (**Supplementary Figure 2**), ranging from 20-40% for most of the past two decades, compared to 40-50% in adults. However, in 2014-15, the percentage notably increased in children to the same level as adults, reaching above 50% for both.

**Figure 4.**
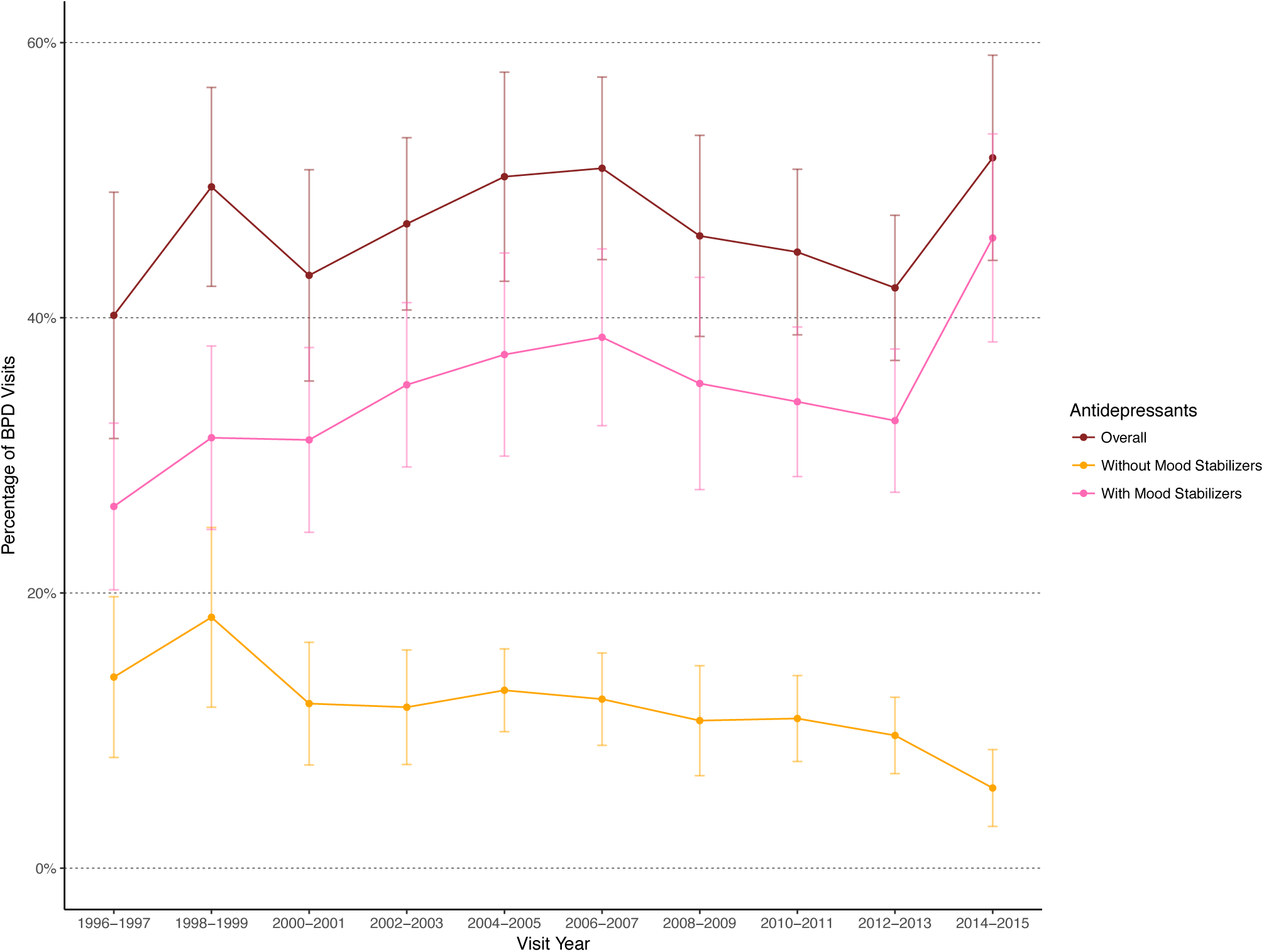
Trends in prescriptions of antidepressants for BPD ^*i*^. *i* Error bars show for each weighted estimate of the percentage of BPD visits the 95% confidence interval.

## Discussion

Lithium, the first medication identified to be effective for treating BPD, remains a first line therapy (Severus et al., 2012). However, the use of lithium has been steadily declining over the past thirty years. The results reported in this paper further document the decline in prescriptions of lithium to approximately 15% of physician office visits for BPD in the US. In its place, there has been a notable increase in prescriptions of mood stabilizing anticonvulsants, initially valproic acid, followed by lamotrigine, and more recently the SGAs, particularly quetiapine and aripiprazole. Mood stabilizing anticonvulsants and SGAs are each now prescribed in approximately 50% of BPD visits, and they are concomitantly prescribed in over 20% of these visits. Moreover, we found that polytherapy with mood stabilizing treatments for BPD has steadily increased since 1998-99, such that two or more of any mood stabilizer medications are now prescribed in over 35% of BPD visits. By contrast, prescriptions of lithium monotherapy have steadily declined over this time period to approximately 3.5% of BPD visits.

The current findings are consistent with those from previous studies in the US. An earlier analysis of the NAMCS data using only outpatient psychiatrist visits found that prescriptions of lithium declined from 50.9% (95%CI: 47.0% - 54.8%) in 1992-95 to 30.1% (95%CI: 26.5% - 33.7%) in 1996-99 (Blanco et al., 2002). Our findings extend this observation into the first two decades of the 2000’s. In addition, a smaller study of patients referred to a tertiary care referral clinic found that lamotrigine, quetiapine, and aripiprazole use more than doubled from 2000-05 to 2006-11, while lithium, valproate, olanzapine and risperidone use decreased (Hooshmand et al., 2014). Our results show similar trends at the national level.

Interestingly, studies from other countries have suggested a constant or even an increasing rate of lithium use (Bramness et al., 2009; Castells et al., 2006; Hayes et al., 2011; Wilting et al., 2008). Many of those studies, however, covered either an earlier (all before 2009) or a shorter time frame, which might partly explain the differences between their findings and our study.

Our study used data from a nationally representative sample of office based clinical visits across the US. In addition, these data covered a twenty-year period from 1996-2015 and were collected using largely consistent procedures. This allowed us to have a relatively large sample to make statistical inferences, which is an advantage over past BPD studies. By combining the data into two-year intervals, we attempted to obtain more stable results while not losing the ability to observe time trends. Nevertheless, the study sample may not have been sufficiently large to detect trends in subgroups of patients or trends specific to individual medications. Furthermore, the cross-sectional nature of NAMCS data did not allow assessment of prescribing patterns within individuals longitudinally.

Despite these limitations, the declining use of lithium that we observed in the US is notable. Lithium has been shown to be an effective treatment for BPD, but it is not effective in all patients (M. Gitlin, 2016). Although the introduction of mood stabilizing anticonvulsants and SGAs have provided alternative treatments for BPD, there is limited evidence regarding the comparative safety and effectiveness of these medications. As a result, BPD patients often undergo a long period of trial and error before finding the right treatment. The increasing polytherapy with mood stabilizers that we observed has also been documented in BPD samples from other countries and in psychiatry visits overall (Hayes et al., 2011; Mojtabai & Olfson, 2010). This is of interest because there may be even less empirical evidence to guide which combinations of mood stabilizers are most effective despite increased risk for side effects. Since there is limited evidence to help guide which of these options will work best for different patients, further research is needed to achieve the promise of precision medicine to better predict which medications or medication combinations are most effective for individual patients, thus minimizing the typical process of trial and error of finding the right medications.

## Data Availability

NAMCS data are publicly available through CDC.

## Acknowledgement

We thank Kira E. Riehm and Elizabeth J. Letourneau for their suggestions and comments.

---

**Supplementary Figure 1.**
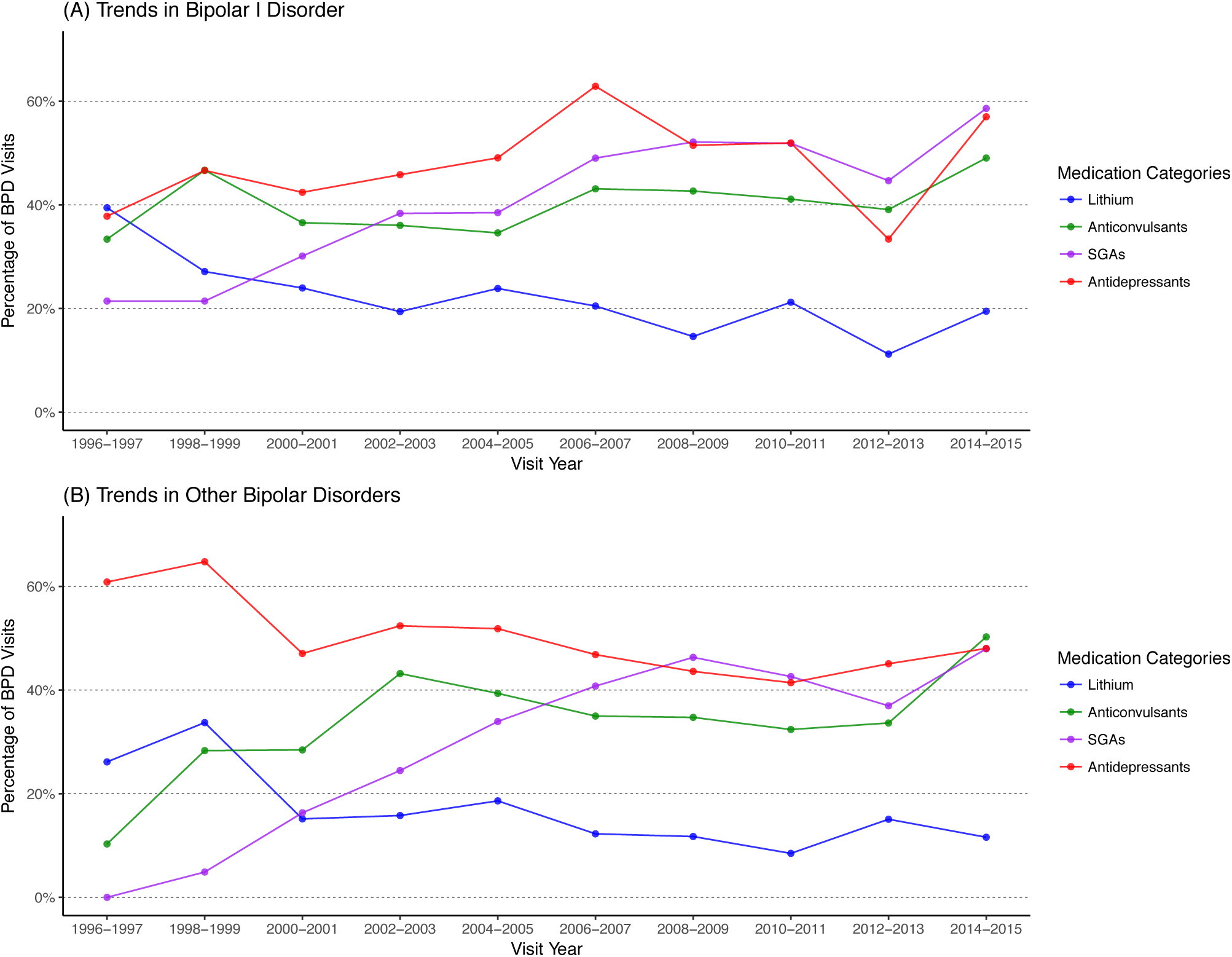
Trends between 1996 and 2015 in the percentage of BPD visits for different categories of medications for bipolar I disorder versus other bipolar disorders^*i*^. *i* Error bars are not shown due to smaller sample sizes of the stratified results.

**Supplementary Figure 2.**
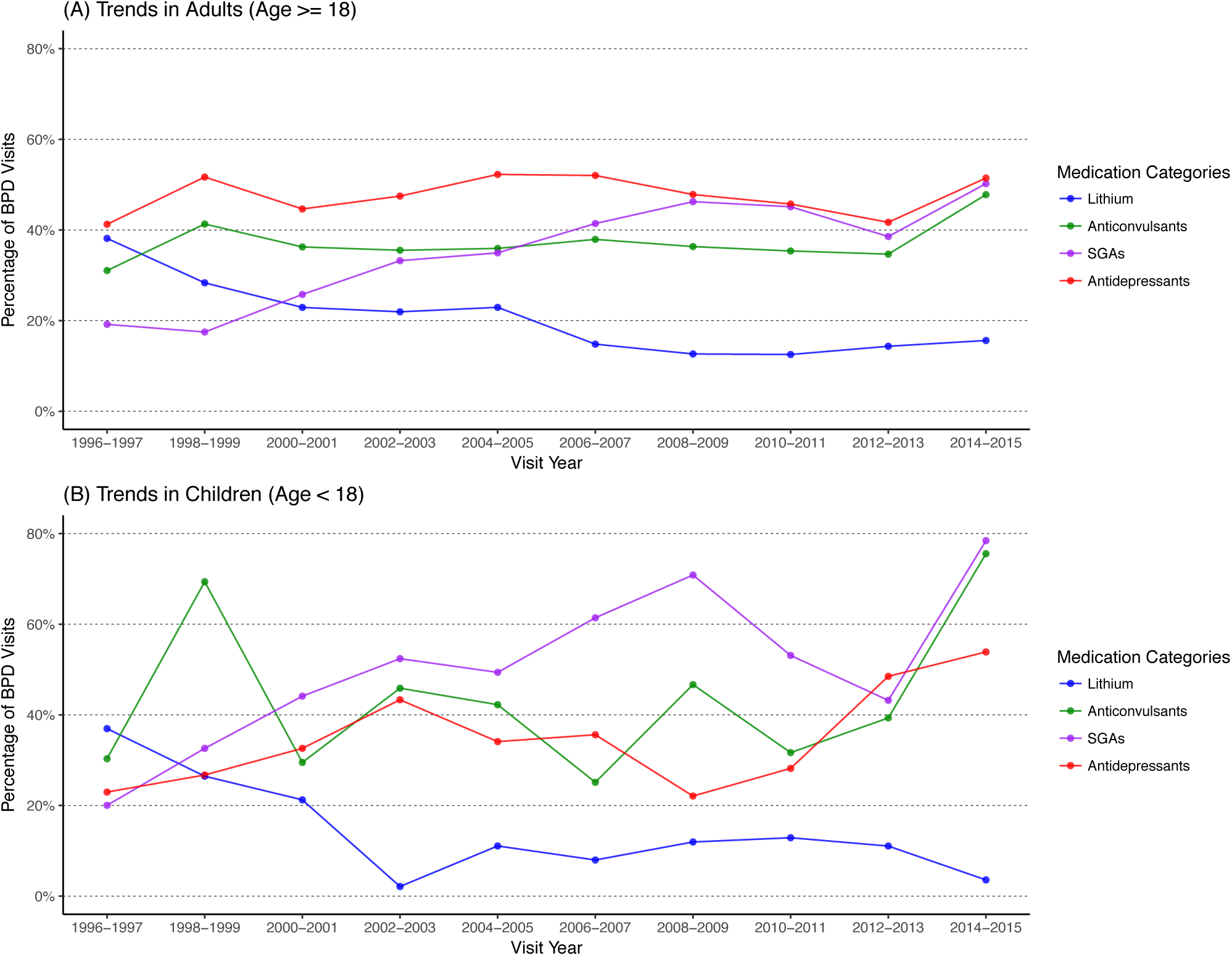
Trends between 1996 and 2015 in the percentage of BPD visits for different categories of medications for BPD in adults versus children^*i*^. *i* Error bars are not shown due to smaller sample sizes of the stratified results.

## Reference

Blanco, C., Laje, G., Olfson, M., Marcus, S. C., & Pincus, H. A. (2002). Trends in the treatment of bipolar disorder by outpatient psychiatrists. Am J Psychiatry, 159(6), 1005–1010. doi: 10.1176/appi.ajp.159.6.1005

Bramness, J. G., Weitoft, G. R., & Hallas, J. (2009). Use of lithium in the adult populations of Denmark, Norway and Sweden. J Affect Disord, 118(1-3), 224–228. doi: 10.1016/j.jad.2009.01.024

Cade, J. F. (1949). Lithium salts in the treatment of psychotic excitement. Med J Aust, 2(10), 349–352.

Caley, C. F., Perriello, E., & Golden, J. (2018). Antiepileptic drugs and suicide-related outcomes in bipolar disorder: A descriptive review of published data. Ment Health Clin, 8(3), 138–147. doi: 10.9740/mhc.2018.05.138

Castells, X., Vallano, A., Rigau, D., Perez, J., Casas, M., & Capella, D. (2006). Trends in lithium prescription in Spain from 1985 to 2003. J Affect Disord, 91(2-3), 273–276. doi: 10.1016/j.jad.2006.01.005

CDC. (2015a). NAMCS scope and sample design. from https://www.cdc.gov/nchs/ahcd/ahcd_scope.htm

CDC. (2015b). Trend analysis using NAMCS and NHAMCS drug data. from https://www.cdc.gov/nchs/ahcd/trend_analysis.htm

CDC. (2017). National Ambulatory Medical Care Survey. from https://www.cdc.gov/nchs/ahcd/about_ahcd.htm

Chesney, E., Goodwin, G. M., & Fazel, S. (2014). Risks of all-cause and suicide mortality in mental disorders: a meta-review. World Psychiatry, 13(2), 153–160. doi: 10.1002/wps.20128

Geddes, J. R., Calabrese, J. R., & Goodwin, G. M. (2009). Lamotrigine for treatment of bipolar depression: independent meta-analysis and meta-regression of individual patient data from five randomised trials. Br J Psychiatry, 194(1), 4–9. doi: 10.1192/bjp.bp.107.048504

Geoffroy, P. A., Etain, B., Henry, C., & Bellivier, F. (2012). Combination therapy for manic phases: a critical review of a common practice. CNS Neurosci Ther, 18(12), 957–964. doi: 10.1111/cns.12017

Gitlin, M. (2016). Lithium side effects and toxicity: prevalence and management strategies. Int J Bipolar Disord, 4(1), 27. doi: 10.1186/s40345-016-0068-y

Gitlin, M. J. (2018). Antidepressants in bipolar depression: an enduring controversy. Int J Bipolar Disord, 6(1), 25. doi: 10.1186/s40345-018-0133-9

Hayes, J., Prah, P., Nazareth, I., King, M., Walters, K., Petersen, I., & Osborn, D. (2011). Prescribing trends in bipolar disorder: cohort study in the United Kingdom THIN primary care database 1995-2009. PLoS One, 6(12), e28725. doi: 10.1371/journal.pone.0028725

Hooshmand, F., Miller, S., Dore, J., Wang, P. W., Hill, S. J., Portillo, N., & Ketter, T. A. (2014). Trends in pharmacotherapy in patients referred to a bipolar specialty clinic, 2000-2011. J Affect Disord, 155, 283–287. doi: 10.1016/j.jad.2013.10.054

Hou, L., Heilbronner, U., Degenhardt, F., Adli, M., Akiyama, K., Akula, N., … Schulze, T. G. (2016). Genetic variants associated with response to lithium treatment in bipolar disorder: a genome-wide association study. Lancet, 387(10023), 1085–1093. doi: 10.1016/S0140-6736(16)00143-4

Hunkeler, E. M., Fireman, B., Lee, J., Diamond, R., Hamilton, J., He, C. X., … Hargreaves, W. A. (2005). Trends in use of antidepressants, lithium, and anticonvulsants in Kaiser Permanente-insured youths, 1994-2003. J Child Adolesc Psychopharmacol, 15(1), 26–37. doi: 10.1089/cap.2005.15.26

Kessing, L. V., Sondergard, L., Kvist, K., & Andersen, P. K. (2005). Suicide risk in patients treated with lithium. Arch Gen Psychiatry, 62(8), 860–866. doi: 10.1001/archpsyc.62.8.860

Lin, D., Mok, H., & Yatham, L. N. (2006). Polytherapy in bipolar disorder. CNS Drugs, 20(1), 29–42. doi: 10.2165/00023210-200620010-00003

Lindstrom, L., Lindstrom, E., Nilsson, M., & Hoistad, M. (2017). Maintenance therapy with second generation antipsychotics for bipolar disorder - A systematic review and meta-analysis. J Affect Disord, 213, 138–150. doi: 10.1016/j.jad.2017.02.012

Manchia, M., Adli, M., Akula, N., Ardau, R., Aubry, J. M., Backlund, L., … Alda, M. (2013). Assessment of Response to Lithium Maintenance Treatment in Bipolar Disorder: A Consortium on Lithium Genetics (ConLiGen) Report. PLoS One, 8(6), e65636. doi: 10.1371/journal.pone.0065636

McKnight, R. F., Adida, M., Budge, K., Stockton, S., Goodwin, G. M., & Geddes, J. R. (2012). Lithium toxicity profile: a systematic review and meta-analysis. Lancet, 379(9817), 721–728. doi: 10.1016/S0140-6736(11)61516-X

Merikangas, K. R., Akiskal, H. S., Angst, J., Greenberg, P. E., Hirschfeld, R. M., Petukhova, M., & Kessler, R. C. (2007). Lifetime and 12-month prevalence of bipolar spectrum disorder in the National Comorbidity Survey replication. Arch Gen Psychiatry, 64(5), 543–552. doi: 10.1001/archpsyc.64.5.543

Merikangas, K. R., Jin, R., He, J. P., Kessler, R. C., Lee, S., Sampson, N. A., … Zarkov, Z. (2011). Prevalence and correlates of bipolar spectrum disorder in the world mental health survey initiative. Arch Gen Psychiatry, 68(3), 241–251. doi: 10.1001/archgenpsychiatry.2011.12

Mohr, P. (2007). Quality of life in the long-term treatment and the role of second-generation antipsychotics. Neuro Endocrinol Lett, 28 Suppl 1, 117–133.

Mojtabai, R., & Olfson, M. (2010). National trends in psychotropic medication polypharmacy in office-based psychiatry. Arch Gen Psychiatry, 67(1), 26–36. doi: 10.1001/archgenpsychiatry.2009.175

Moreno, C., Laje, G., Blanco, C., Jiang, H., Schmidt, A. B., & Olfson, M. (2007). National trends in the outpatient diagnosis and treatment of bipolar disorder in youth. Arch Gen Psychiatry, 64(9), 1032–1039. doi: 10.1001/archpsyc.64.9.1032

Nierenberg, A. A., Friedman, E. S., Bowden, C. L., Sylvia, L. G., Thase, M. E., Ketter, T., … Calabrese, J. R. (2013). Lithium treatment moderate-dose use study (LiTMUS) for bipolar disorder: a randomized comparative effectiveness trial of optimized personalized treatment with and without lithium. Am J Psychiatry, 170(1), 102–110. doi: 10.1176/appi.ajp.2012.12060751

Okusa, M. D., & Crystal, L. J. (1994). Clinical manifestations and management of acute lithium intoxication. Am J Med, 97(4), 383–389.

Sachs, G. S., Nierenberg, A. A., Calabrese, J. R., Marangell, L. B., Wisniewski, S. R., Gyulai, L., … Thase, M. E. (2007). Effectiveness of adjunctive antidepressant treatment for bipolar depression. N Engl J Med, 356(17), 1711–1722. doi: 10.1056/NEJMoa064135

Severus, E., Schaaff, N., & Moller, H. J. (2012). State of the art: treatment of bipolar disorders. CNS Neurosci Ther, 18(3), 214–218. doi: 10.1111/j.1755-5949.2011.00258.x

Weisler, R. H., Cutler, A. J., Ballenger, J. C., Post, R. M., & Ketter, T. A. (2006). The use of antiepileptic drugs in bipolar disorders: a review based on evidence from controlled trials. CNS Spectr, 11(10), 788–799.

Wilting, I., Souverein, P. C., Nolen, W. A., Egberts, A. C., & Heerdink, E. R. (2008). Changes in outpatient lithium treatment in the Netherlands during 1996-2005. J Affect Disord, 111(1), 94–99. doi: 10.1016/j.jad.2008.01.019

